# AI-enabled Left Atrial Volumetry in Cardiac CT Scans Improves CHARGE-AF and Outperforms NT-ProBNP for Prediction of Atrial Fibrillation in Asymptomatic Individuals: Multi-Ethnic Study of Atherosclerosis

**DOI:** 10.1101/2024.01.22.24301384

**Authors:** Morteza Naghavi, David Yankelevitz, Anthony P. Reeves, Matthew J. Budoff, Dong Li, Kyle C. Atlas, Chenyu Zhang, Thomas L. Atlas, Seth Lirette, Jakob Wasserthal, Claudia Henschke, Christopher Defilippi, Susan R. Heckbert, Philip Greenland

**Author notes:** **Address for correspondence:** Morteza Naghavi, 450 Holcombe, Houston, TX, 77021.

## Abstract

**Background:** Coronary artery calcium (CAC) scans contain actionable information beyond CAC scores that is not currently reported.

**Methods:** We have applied artificial intelligence-enabled automated cardiac chambers volumetry to CAC scans (AI-CAC), taking on average 21 seconds per CAC scan, to 5535 asymptomatic individuals (52.2% women, ages 45-84) that were previously obtained for CAC scoring in the baseline examination (2000-2002) of the Multi-Ethnic Study of Atherosclerosis (MESA). We used the 5-year outcomes data for incident atrial fibrillation (AF) and compared the time-dependent AUC of AI-CAC LA volume with known predictors of AF, the CHARGE-AF Risk Score and NT-proBNP (BNP). The mean follow-up time to an AF event was 2.9±1.4 years.

**Results:** At 1,2,3,4, and 5 years follow-up 36, 77, 123, 182, and 236 cases of AF were identified, respectively. The AUC for AI-CAC LA volume was significantly higher than CHARGE-AF or BNP at year 1 (0.836, 0.742, 0.742), year 2 (0.842, 0.807,0.772), and year 3 (0.811, 0.785, 0.745) (p<0.02), but similar for year 4 (0.785, 0.769, 0.725) and year 5 (0.781, 0.767, 0.734) respectively (p>0.05). AI-CAC LA volume significantly improved the continuous Net Reclassification Index for prediction of AF over years 1-5 when added to CAC score (0.74, 0.49, 0.53, 0.39, 0.44), CHARGE-AF Risk Score (0.60, 0.28, 0.32, 0.19, 0.24), and BNP (0.68, 0.44, 0.42, 0.30, 0.37) respectively (p<0.01).

**Conclusion:** AI-CAC LA volume enabled prediction of AF as early as one year and significantly improved on risk classification of CHARGE-AF Risk Score and BNP.

## Introduction

Coronary artery calcium (CAC) scoring is the strongest predictor of atherosclerotic cardiovascular disease (ASCVD) in asymptomatic individuals available today^1^. However, it is a weak predictor of atrial fibrillation (AF), the most common sustained arrhythmia that significantly increases the risk of stroke and cardiovascular mortality^2^. Incident AF is on the rise leading to morbidity and mortality worldwide, both in the elderly and among younger adults^2,3,4,5,6,7^. Currently for prediction of AF in asymptomatic population we are limited to the CHARGE-AF Risk Score which is an epidemiological risk calculator created based on both asymptomatic people and patients with cardiovascular disease (CVD). Amino terminal Pro-B-type natriuretic peptide (NT-proBNP) is a blood protein that is associated with enlarged cardiac chambers and correlates with left atrial (LA) volume. Recent studies have linked NT-proBNP to the incidence of AF and reported incremental predictive value when BNP is added to the CHARGE-AF Risk Score^8,9^.

Since left atrial diameter and strain are known to be associated with risk for developing atrial fibrillation^10,11^, and pioneering efforts from Heinz Nixdorf Recall Study showed the potential value of non-coronary findings in CAC scans^12,13,14,15,16^, we hypothesized that AI-powered cardiac chambers volumetry in CAC scans (AI-CAC) could enable AF prediction in asymptomatic individuals. In this study, we present AI-CAC data obtained from existing CAC scans in a large prospective study and compare the predictive value of AI-CAC estimated LA volume versus the CHARGE-AF Score and BNP for predicting AF.

## Methods

### Study population

The Multi-Ethnic Study of Atherosclerosis (MESA) is a prospective, population-based, observational cohort study of 6,814 men and women without clinical cardiovascular disease (CVD) at the time of recruitment from six field centers in the United States. As part of the initial evaluation (2000-2002), participants received a comprehensive medical history, clinic examination, and laboratory tests. Demographic information, medical history, and medication use at baseline were obtained by self-report. An ECG-gated non-contrast CT was performed at the baseline examination to measure CAC. Non-CT scan covariates included BNP and variables used in calculating the CHARGE-AF Risk Score. Details on BNP assays measurements are described below under BNP Measurement. Covariates used in CHARGE-AF Score for our analyses are age, gender, ethnicity, height, weight, systolic blood pressure, diastolic blood pressure, current smoking, hypertension medication, diabetes, which were obtained as a part of MESA baseline exam 1 previously described^18^. Additionally, CHARGE-AF Risk Score includes myocardial infarction and heart failure which were by default absent in the asymptomatic MESA population at baseline exam 1.

For our study, we removed 771 MESA participants who did not consent for commercial use of data, leaving 6043 participants for our analysis. After removing 125 cases with missing slices in CAC scans, 4 cases with missing data for CHARGE-AF Risk Score, and 168 cases with missing BNP values we have 5746 remaining participants. Subsequently, we have removed 70 cases with pre-baseline AF, 9 cases with surgical AF, and 132 non-AF deaths resulting in the total number of 5535 cases available for analysis.

### Outcomes

Participants were contacted by telephone every 9-12 months during follow-up and asked to report all new cardiovascular diagnoses. International Classification of Disease (ICD) codes were obtained. Incident AF was identified by ICD codes 427.3x (version 9) or I48.x (version 10) from inpatient stays and, for participants enrolled in fee-for-service Medicare, from Medicare claims for outpatient and provider services. For participant reports of heart failure, coronary heart disease, stroke, and CVD mortality, detailed medical records were obtained, and diagnoses were adjudicated by the MESA Morbidity and Mortality Committee. Additionally, BNP data was obtained from MESA core laboratory for MESA exam 1 participants. A detailed study design for MESA has been published elsewhere^18^. MESA participants have been followed since the year 2000. Incident AF has been identified through December 2018. 70 cases with previously diagnosed AF prior to MESA enrollment were removed from the analysis.

### The AI tool for Automated Cardiac Chambers Volumetry

The automated cardiac chambers volumetry tool referred to in this study is called AutoChamber^TM^ (HeartLung.AI, Houston, TX), a deep learning model that used TotalSegmentator^19^ as the base input and was further developed to segment not only each of the four cardiac chambers; left atrium (LA), left ventricle (LV), right atrium (RA), and right ventricle (RV) but also ascending aorta, aortic root and valves, pulmonary arteries, and several other components which are not presented here. The AI-CAC LA volumetry is the focus of this manuscript. Figure 1 shows the segmentations of cardiac chambers in color. The base architecture of the TotalSegmentator model was trained on 1139 cases with 447 cases of coronary CT angiography (CCTA) using nnU-Net, a self-configuring method for deep learning-based biomedical image segmentation^20^. The initial input training data were matched non-contrast and contrast-enhanced ECG-gated cardiac CT scans with 1.5 mm slice thickness.

**Figure 1.**
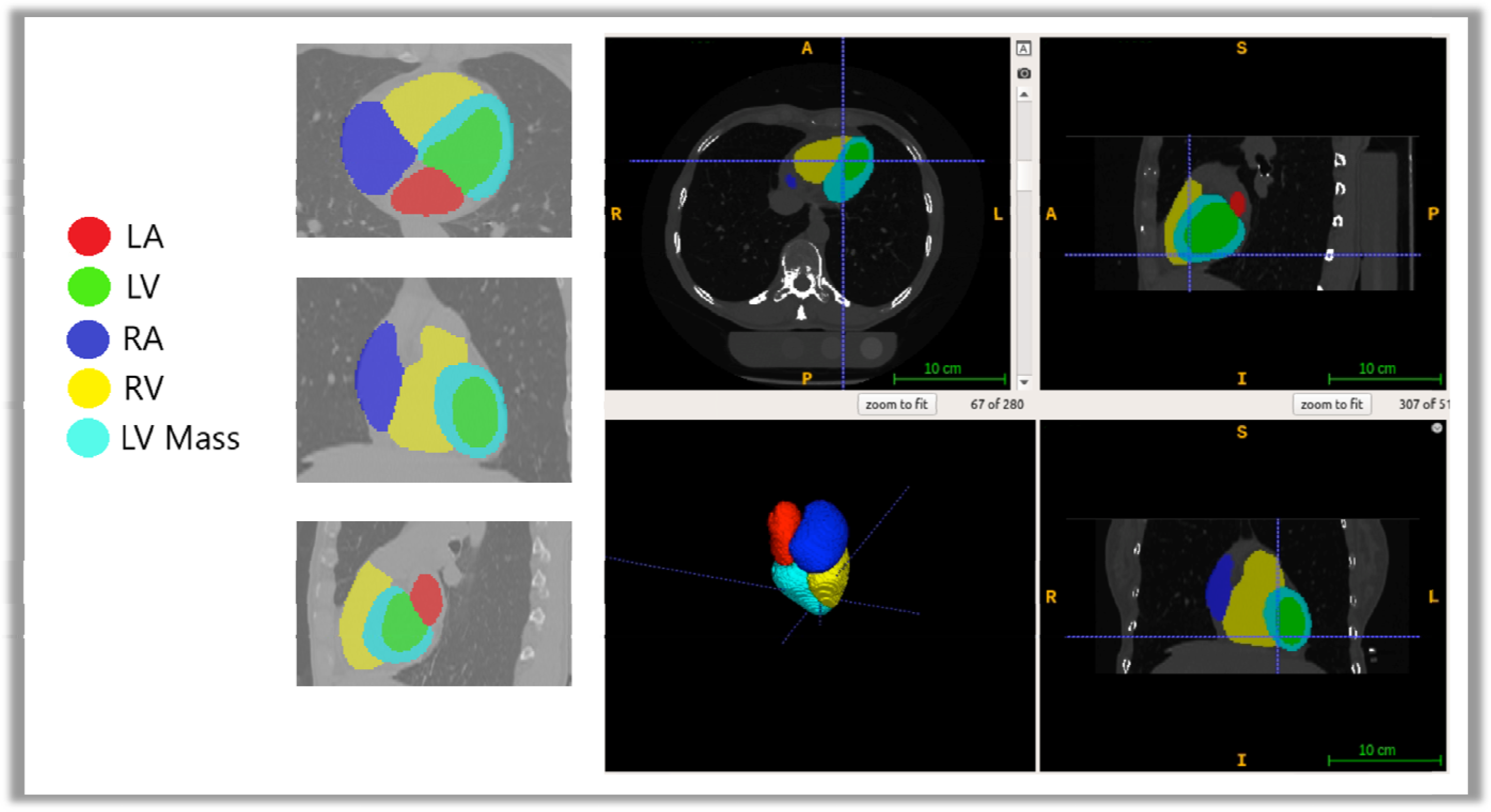
Examples of AI-CAC segmentations in a cardiac CT scan.

Because the images were taken from the same patients in the same session, registration was done with good alignment. Following this transfer of segmentations, a nnU-Net deep learning tool was used for training the model. Additionally, iterative training was implemented whereby human supervisors corrected errors made by the model, and the corrected data were used to further train the model, leading to improved accuracy. To standardize the comparison in MESA, cardiac chambers were reported by gender and ethnicity adjusted by body surface area (BSA) using residual adjustment techniques. (BSA: 0.007184 x (height(m)^0.725) x (weight(kg)^0.425)).

Additionally, an internal reference was developed based on the field of view size and the posterior height of thoracic vertebral bones. This measure would be used whenever BSA information is unavailable, however it was not an issue in MESA. AutoChamber™ AI was run on 6043 non-contrast CAC scans that consented to commercial data usage out of the 6814 scans available in MESA exam 1. Expert rules built in the AI-model excluded 125 cases due to missing slices in image reconstruction created by some of the electron beam CT scanners used in MESA baseline. These cases were random, and our investigations did not reveal any particular association with dependent or independent variables in our study (see Results).

### CHARGE-AF Risk Score

The CHARGE-AF risk score was developed to predict risk of incident AF in three American cohorts, and it was validated in two European cohorts. The linear predictor from the CHARGE-AF Risk Score is calculated as: (age in years/5) * 0.5083+ethnicity (Caucasian/white) * 0.46491 + (height in centimeters/10) * 0.2478 + (weight in kg/15) * 0.1155 + (SBP in mm Hg/20) * 0.1972 – (DBP in mm Hg/10) * 0.1013+current smoking * 0.35931+antihypertensive medication use * 0.34889+DM * 0.23666^21^. The result is the sum of the product of the regression coefficients and the predictor variables, which represents the change in the hazard ratio for a one-unit change in the corresponding predictor variable.

### BNP Measurement

Details on BNP assays used in MESA have been reported^17^. N-terminal proBNP is more reproducible than BNP at the lower end of the distribution range, and more stable at room temperature. However, both BNP and N-terminal proBNP are clinically available. Intra-assay and inter-assay coefficients of variation at various concentrations of NT-proBNP have been previously reported^22,23^. The analytical measurement range for NT-proBNP in exam 1 was 4.9– 11699 pg/ml. The lower limits of detection for the NT-proBNP assay is 5 pg/mL, thus cases above 0 and below 4.99 were treated as 4.99 pg/mL. Clinically, values are not reported below 4.99 pg/mL because the analytical accuracy is poor at those low levels (i.e. typically a coefficient of variation of greater than 20% between repeat measures).

### Statistical Analysis

We used SAS (SAS Institute Inc., Cary, NC) and Stata (StatCorp LLC, College Station, TX) software for statistical analyses. All values are reported as means ± SD except for BNP which did not show normal distribution and is presented in median and interquartile range (IQR). All tests of significance were two tailed, and significance was defined at the p<0.05 level. Cumulative incidence was calculated using one minus the Kaplan-Meier survival estimate. Group differences in incidence were determined using the log-rank test.

Cox proportional hazards regression was used for survival analysis. The time-dependent ROC (receiver operator curve) AUC (area under the curve) was calculated using the inverse probability of censoring weighting estimator. Hazard ratios over 5 years were calculated per SD. BNP and CAC were natural logarithm-transformed (ln-transformed) to avoid undue influence of large values. AI-CAC LA volume and CHARGE-AF Risk Score showed a normal distribution.

Category-free (continuous) net reclassification index (NRI) was calculated using the sum of the differences between the proportions of upward reclassifications and downward reclassifications for AF events and AF non-events, respectively. NRI was developed as a statistical measure to evaluate the improvement in risk prediction models when additional variables are incorporated into a base model^24^. We have analyzed data for AF prediction at 1 to 5 years follow up.

### Ethical Approval

This study has received proper ethical oversight. All subjects gave their informed consent for inclusion before they participated in the study. Subjects who did not consent were removed from the study.

## Results

In the cohort, ages ranged from 45-84, 52.2% were women, 39.7% were White, 26.1% Black, 22% Hispanic, and 12.1% Chinese. Table 1 shows the baseline characteristics of MESA participants who were diagnosed with incident AF versus those who were not over the period of 5 years follow up. At 1,2,3,4, and 5 years follow up 36, 77, 123, 182, and 236 cases of AF were identified respectively. In univariate comparisons, incident AF cases were older, more likely male, and more likely White. The incident AF cases had higher cardiac chamber volumes for LA, LV, RA, LV Wall, CHARGE-AF Risk Scores and NT-proBNP levels versus those without incident AF (all comparisons p< 0.001) (Table 1).

**Table 1:** Baseline characteristics of the Multi-Ethnic Study of Atherosclerosis (MESA) participants including cases with and without Atrial Fibrillation (AF) at 5 years.

|  | Overall<br>(N=5535) | No AF†<br>(N=5319) | AF*<br>(N=216) | P<br>Value |
| --- | --- | --- | --- | --- |
| **Age (per 10 years)** |  |  |  |  |
| Age 45-54 | 28.9% | 29.7% | 2.1% | <.0001 |
| Age 55-64 | 27.6% | 28.0% | 18.1% | <.0001 |
| Age 65-74 | 29.4% | 28.8% | 46.1% | <.0001 |
| Age 75-84 | 14.1% | 13.5% | 33.7% | <.0001 |
| Female sex (%) | 52.2% | 52.8% | 47.9% | <.0001 |
| Body Surface Area | 1.90±0.24 | 1.89±0.24 | 1.92±0.25 | <.0001 |
| **Ethnicity** |  |  |  |  |
| White | 39.5% | 46.6% | 39.4% | 0.0229 |
| Chinese | 12.4% | 11.4% | 12.2% | 0.7054 |
| Black | 25.8% | 21.2% | 26.3% | 0.0615 |
| Hispanic | 22.3% | 20.8% | 22% | 0.6569 |

| **AI-Enabled Cardiac Chambers Volumetry** |  |  |  |  |
| --- | --- | --- | --- | --- |
| LV volume (mm<sup>3</sup>) | 102.23±24.96 | 102.1±25.0 | 108.0±31.1 | <.0001 |
| LA volume (mm<sup>3</sup>) | 60.94±15.10 | 60.6±15.3 | 73.5±24.5 | <.0001 |
| RV volume (mm<sup>3</sup>) | 134.30±34.43 | 134.1±34.4 | 136.0±37.7 | 0.4081 |
| RA volume (mm<sup>3</sup>) | 76.76±18.75 | 76.6±18.4 | 83.3±26.0 | <.0001 |
| LV Wall volume (g) | 107.53±26.08 | 107.3±26.1 | 114.2±30.6 | <.0001 |
| Total heart (mm<sup>3</sup>) | 481.76±108.69 | 480.7±108.1 | 514.9±134.9 | <.0001 |
| CHARGE-AF Score | 11.7±1.2 | 11.7±1.2 | 12.8±0.9 | <.0001 |
| BNP (Median – IQR)‡ | 51.41 (23.19–<br>104.4) | 49.46 (22.54–<br>98.15) | 115.8 (62.42–<br>236) | <.0001 |
| BNP (mean) | 82.1±95.0 | 78.3±89.4 | 175.7±159.6 | <.0001 |
| CAC (Median – IQR)‡ | 0 (0-80.84) | 0 (0-73.34) | 59.52 (3.16-257.60) | <.0001 |
| CAC (mean) | 133.7±379.0 | 125.5±358.8 | 333.3±686.8 | <.0001 |

| **Risk Factors** |  |  |  |  |
| --- | --- | --- | --- | --- |
| Diabetes | 12.1% | 12.1% | 15.7% | 0.0987 |
| Hypertension | 43.8% | 43.4% | 62.7% | <.0001 |
| Smoking (Current use) | 12.8% | 13.0% | 10.6% | 0.2816 |
| Alcohol (Current use) | 69.3% | 69.4% | 63.5% | 0.0547 |
| Blood Pressure Lowering Rx | 36.0% | 35.7% | 54.9% | <.0001 |
| Lipid Lowering Rx | 16.4% | 16.5% | 16.6% | 0.9677 |
| LDL Cholesterol (mg/dl) | 117.2±31 | 117.4±31.2 | 115.4±33.4 | 0.2017 |
| HDL Cholesterol (mg/dl) | 50.9±15 | 51.0±15.0 | 50.0±13.9 | 0.3212 |
| Total Cholesterol (mg/dl) | 194.4±35.3 | 194.5±35.5 | 192.2±38.0 | 0.0021 |

\*AF Events above 5 years are excluded. Mean follow-up years to an AF event 2.9±1.4.
† 132 deaths due to non-AF events are excluded
‡ Only median was used for analysis

The cumulative incidence of AF over 5 years for AI-estimated LA volume, CHARGE-AF Risk Score and BNP are shown in Figure 2. The incidence of AF in the 99^th^ percentile of AI-LA volume, CHARGE-AF Risk Score, and BNP were 37.3%, 16.5%, 27.1 respectively (p<0.0001).

**Figure 2a-d.**
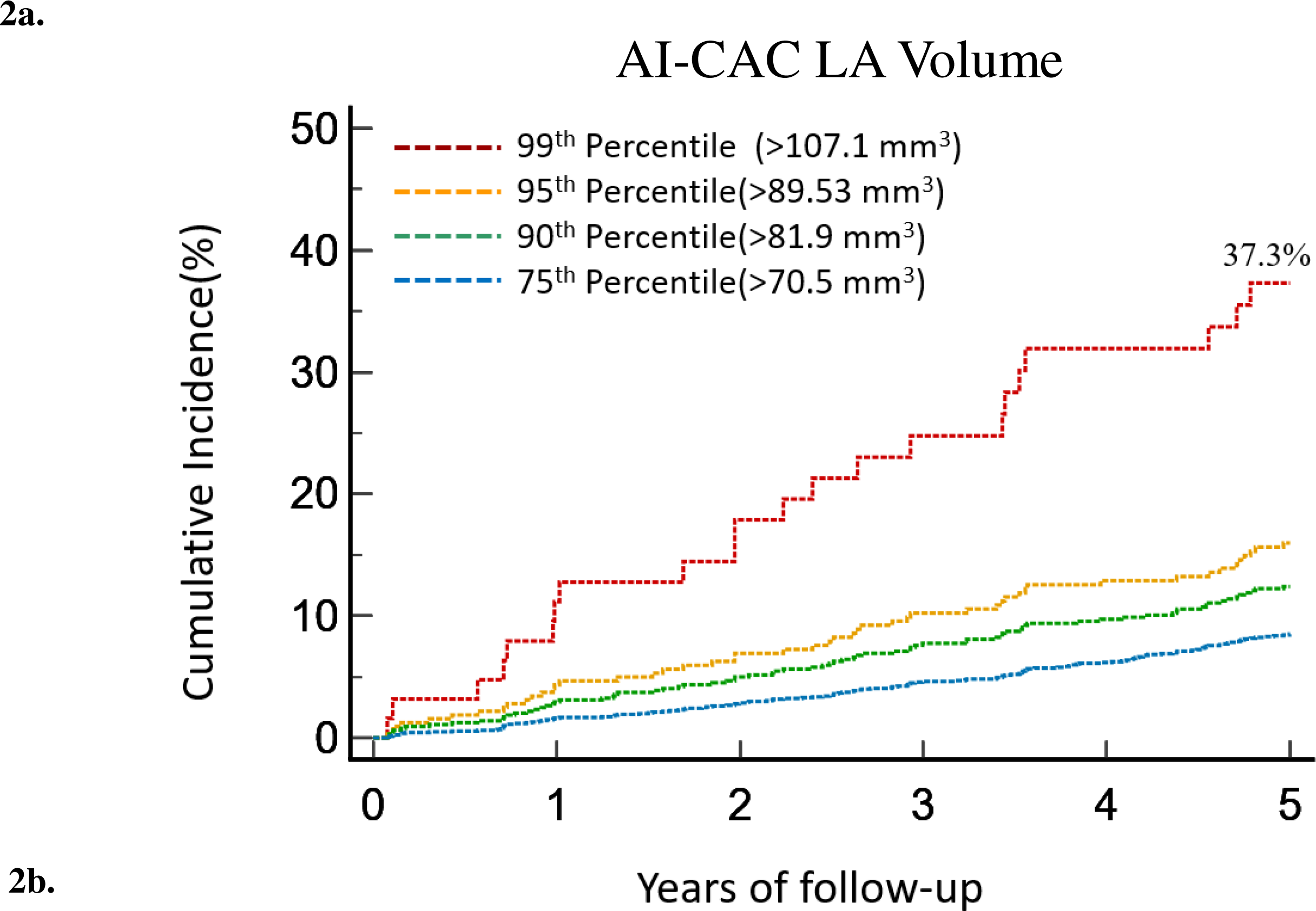
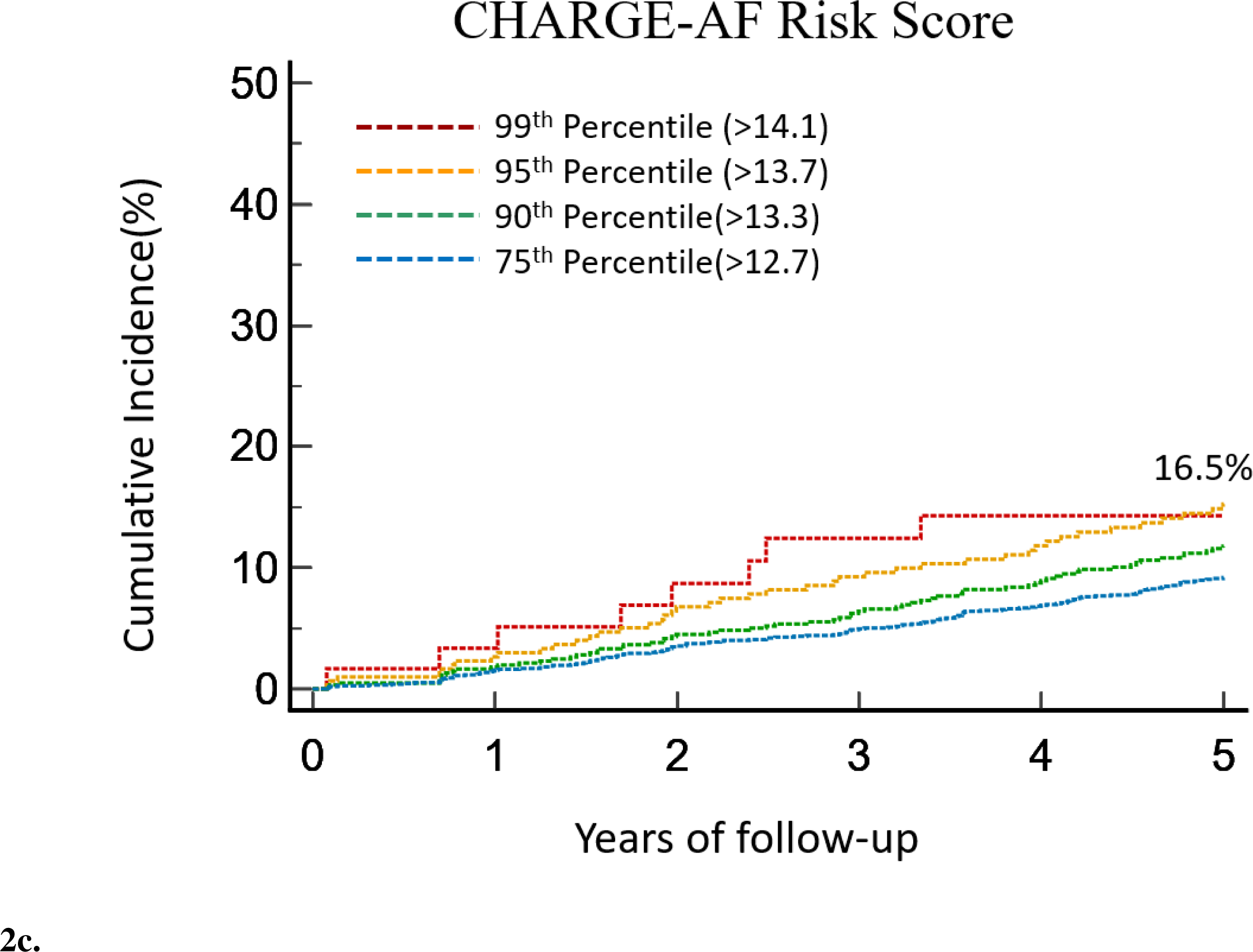
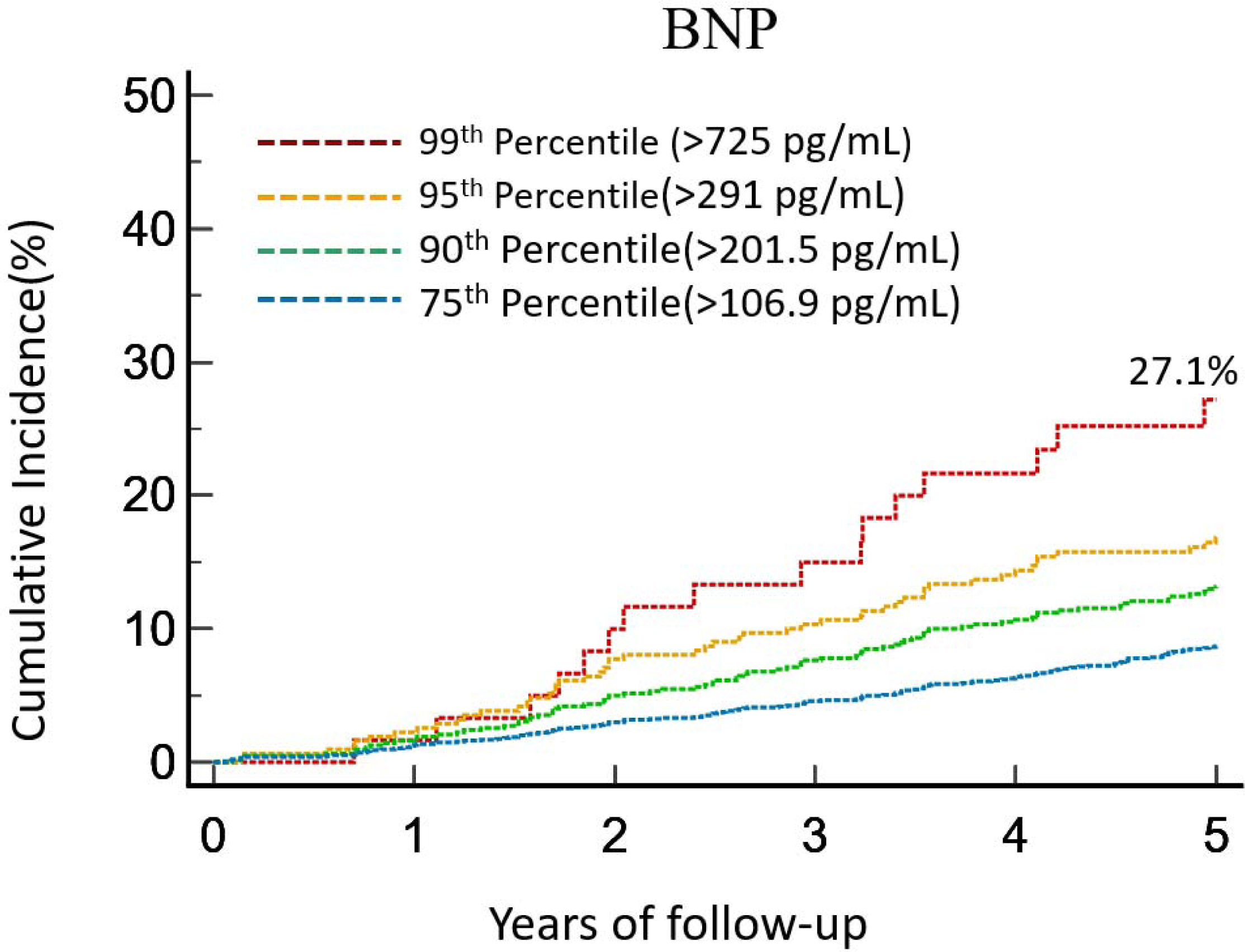
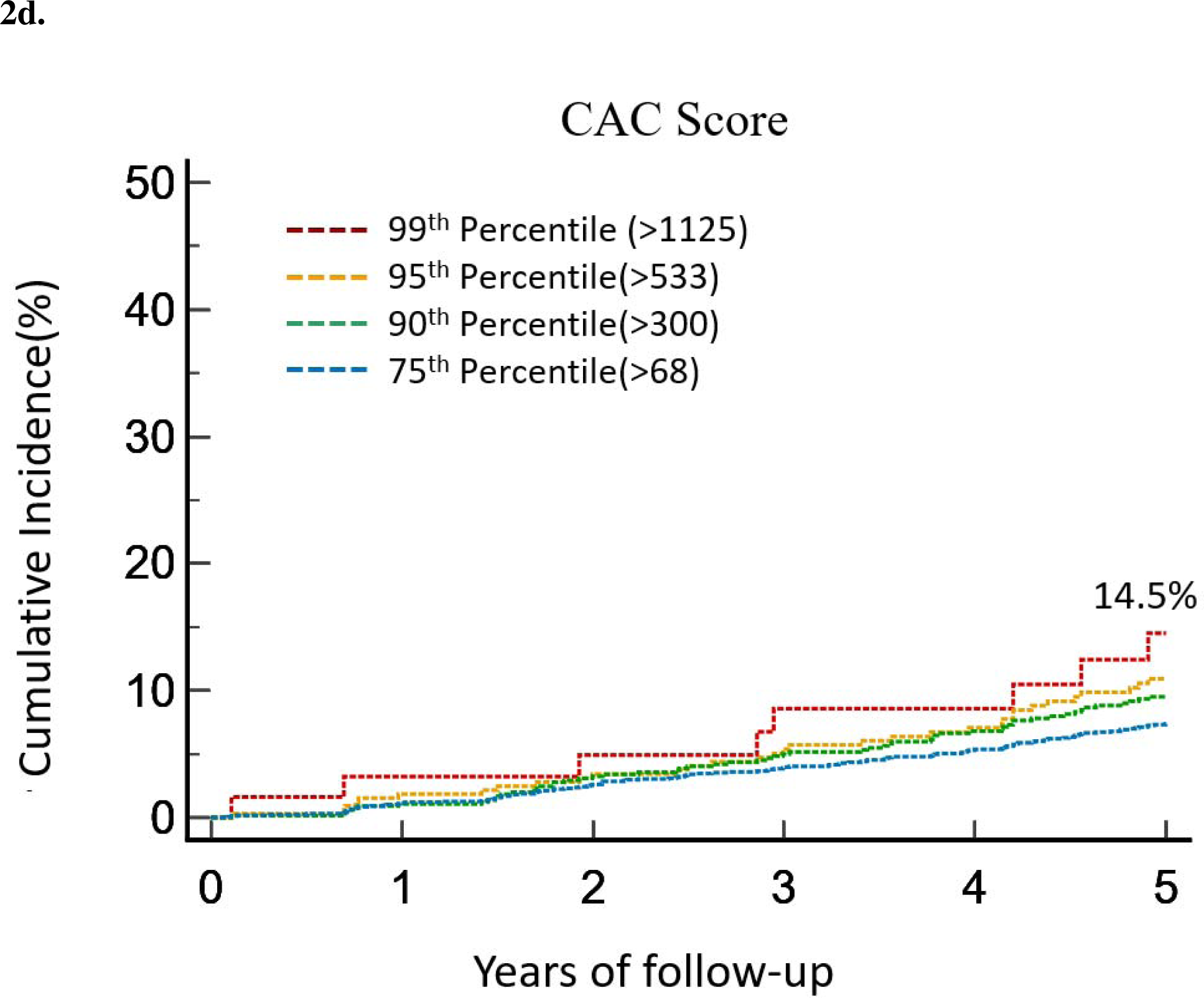
Cumulative Incidence of Atrial Fibrillation (AF) in the Top Quartile of AI-CAC Left Atrial (LA) Volume, CHARGE-AF Score, NT-proBNP (BNP) and coronary artery calcium (CAC) over 5 years of follow-up.

The AUC for AI-estimated LA volume (adjusted by age, gender, BSA) was significantly higher than AUC for CHARGE-AF Risk Score and BNP over 1-5 years (Table 2). When comparing AUC individually between AI-LA volume vs. BNP the differences were statistically significant (p < 0.001) for years 1 to 5. The AUC for AI-LA volume vs. CHARGE AF was statistically significant (p < 0.02) for years 1, 2, 3, but not statistically significant for year 4 (p = 0.11) and year 5 (p=0.08). The difference in AUC for AI-estimated LA volume alone versus CHARGE-AF and BNP combined, despite higher AUC for LA volume in years 1 to 3, was not statistically significant (year 1, 0.836 vs. 0.775, p = 0.07, year 2, 0.842 vs. 0.835, p = 0.66, year 3, 0.811 vs. 0.785, p = 0.99, year 4, 0.785 vs. 0.791, p = 0.50, year 5, 0.781 vs. 0.787, p = 0.41).

**Table 2:** Time-dependent Area Under Curve (AUC) and Net Reclassification Index (NRI) for Atrial Fibrillation (AF) Prediction between AI-CAC Left Atrial (LA) Volume, CHARGE-AF Risk Score, and NT-proBNP (BNP) over 1 to 5 Years in Multi-Ethnic Study of Atherosclerosis (MESA)

|  | 1 Year |  | 2 Years |  | 3 Years |  | 4 Years |  | 5 Years |  |
| --- | --- | --- | --- | --- | --- | --- | --- | --- | --- | --- |
| AF Events | **36** |  | **77** |  | **123** |  | **182** |  | **236** |  |
| **Predictors** | **AUC** | **P value** | **AUC** | **P value** | **AUC** | **P value** | **AUC** | **P value** | **AUC** | **P value** |
| AI-CAC LA Volume* | 0.836 | - | 0.842 | - | 0.811 | - | 0.785 | - | 0.781 | - |
| CHARGE-AF | 0.742 | 0.010 | 0.807 | 0.003 | 0.785 | 0.022 | 0.769 | 0.110 | 0.767 | 0.080 |
| BNP | 0.742 | 0.003 | 0.772 | 0.005 | 0.745 | 0.001 | 0.725 | 0.001 | 0.734 | 0.001 |
| CHARGE-AF + BNP | 0.775 | 0.07 | 0.835 | 0.66 | 0.811 | 0.99 | 0.791 | 0.50 | 0.787 | 0.41 |
| **Category-Free NRI adding AI-LA** | **NRI** | **P value** | **NRI** | **P value** | **NRI** | **P value** | **NRI** | **P value** | **NRI** | **P value** |
| To Base Model CHARGE-AF | 0.60 | <.0001 | 0.28 | <.0001 | 0.33 | <.0001 | 0.19 | <.0001 | 0.23 | <.0001 |
| To Base Model BNP | 0.68 | <.0001 | 0.44 | <.0001 | 0.42 | <.0001 | 0.30 | <.0001 | 0.37 | <.0001 |
| To Base Model CAC | 0.73 | <.0001 | 0.49 | <.0001 | 0.53 | <.0001 | 0.39 | <.0001 | 0.44 | <.0001 |

\*Adjusted by age, gender, and BSA

The continuous NRI for prediction of AF when AI-estimated LA volume was added to CAC score as the only predictor in the base model for years 1-5 were highly significant (0.75, 0.51, 0.53, 0.39, and 0.44 respectively p<0.0001). Similarly, the NRI for AI-LA volume over 1-5 years when added to base model with CHARGE-AF Risk Score (0.60, 0.28, 0.33, 0.19, 0.24) and BNP (0.68, 0.44, 0.42, 0.30, 0.37) were significant (respectively, p for all < 0.0001). (Table 2)

Univariate and multivariate models assessed 5-year HR increase per SD for each predictor for incident AF (Table 3). All predictors were statistically significant in univariate models, while only AI-CAC LA and BNP were significant in multivariate adjustment models based on age, gender, and BSA.

**Table 3:** Five-Year Atrial Fibrillation Risk Prediction: Hazard Ratios per Standard Deviation (SD) Increase in AI-CAC LA Volume, NT-proBNP (BNP), Agatston CAC Score (CAC), and CHARGE-AF Risk Score.

| Predictors | Univariate Model |  |  | Multivariate Model† |  |  |
| --- | --- | --- | --- | --- | --- | --- |
|  | HR (95% CI) | Beta* | P-value | HR (95% CI) | Beta* | P-value |
| AI-CAC LA Volume<br>(per 1 SD) | 1.422 (1.219-1.659) | 0.352 | <.0001 | 1.301 (1.143-1.462) | 0.263 | <.0001 |
| Ln (BNP)<br>(per 1 SD) | 1.306 (1.110-1.545) | 0.267 | <.0001 | 1.288 (1.080-1.568) | 0.253 | 0.0057 |
| Ln (CAC)<br>(per 1 SD) | 1.149 (1.025-1.287) | 0.139 | 0.0153 | 0.992 (0.859-1.146) | -0.008 | 0.9176 |
| CHARGE-AF Score‡<br>(per 1 SD) | 1.464 (1.274-1.683) | 0.381 | <.0001 | - | - | - |

\* Beta per 1 SD increase
† Biomarkers AI-CAC LA Volume, CAC, and BNP adjusted for age, gender, and body surface area (BSA) in a multivariate model.
‡ CHARGE-AF Score could not be adjusted for risk factors similar to biomarkers, as the score is modeled off risk factors.

A considerable portion of participants classified as low-risk for incident AF over 5 years by CHARGE-AF score have enlarged LA (Figure 3).

**Figure 3.**
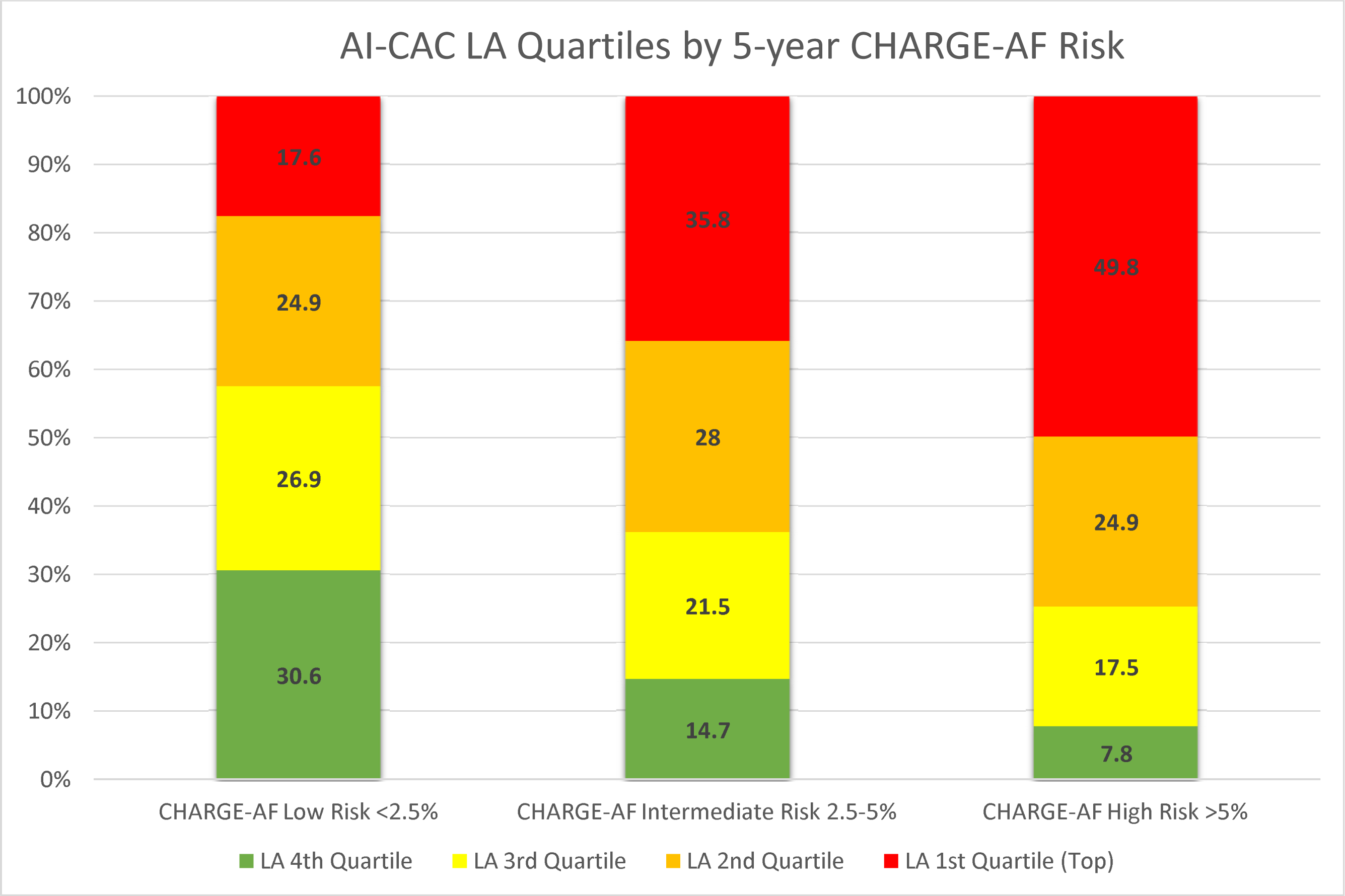
Quartiles of AI-CAC Left Atrial (LA) Volume by predicted 5-year CHARGE-AF Risk

The 125 cases with missing slices were 49.8% male and 50.2% females. None of these cases had a diagnosis of AF. These cases were random, and our investigations did not reveal any association with dependent or independent variables in our study.

## Discussion

To our knowledge this is the first report of an AI-enabled automated cardiac chambers volumetry in non-contrast CT scans obtained for coronary calcium score in a large multi-ethnic study of asymptomatic individuals. Our study demonstrated that the AI-enabled LA volumetry 1) has enabled prediction of AF in CAC scans, 2) significantly outperformed BNP over 1-5 years, 3) significantly outperformed CHARGE-AF Risk Score over 1-3 years, 4) provided for a sizable net reclassification improvement on top of CHARGE-AF Risk Score and BNP, and 5) showed comparable performance against a combined model of CHARGE-AF and BNP over 1-5 years.

CHARGE-AF is an epidemiological risk score for predicting AF based on risk factors at population levels, but it does not lend itself to a useful clinical tool for individualized risk assessment and monitoring of high-risk patients because the large impact of unmodifiable risk factors. For example, if a patient loses 30 lbs. or lowers systolic blood pressure by 20mmHg, the linear predictor from the CHARGE-AF Risk Score only goes down by 0.1. This would be a very minimal change (0.8%) knowing the average CHARGE-AF score in MESA AF cases was 12.8±0.9. Nonetheless, in the absence of an individualized metric with comparable predictive power, it serves as a useful tool for estimating risk and alerting high risk populations to reduce future AF risk^21,25^.

BNP is a serum biomarker of cardiac volume overload particularly and has been studied extensively in various cardiovascular diseases, particularly heart failure^26,27,28^. Thejus et al have shown values above the 80th percentile (97 pg/ml in women and 60 pg/ml in men) present an odds ratio of 2.65 for the incidence of AF^29^. Asselberg et al^30^ found that in the general population, elevated BNP levels at baseline predicted the development of AF when reassessed at 4 years.

The baseline median level was 62.2 pg/ml in those who eventually developed AF compared to 35.7 pg/ml in those who did not (p = 0.001). Our study shows that LA volume outperformed BNP in MESA consistently over 5 and improved its predictive value by NRI of 0.69 for year 1 to NRI of 0.38 for year 5. This may be due to the fact that BNP is not specific to LA or RA volume and can be influenced by other factors.

Although ECG-based screening for AF is currently a topic of great clinical interest^31,32^ it would not be a proper comparison for this study because ECG is primarily used for the detection of prevalent AF not for prediction of future AF. However, recent studies suggest that AI-enabled ECG could play a role in predicting future AF^33,34,35^ .A study by Christopoulos et al that compared the performance of AI-enabled ECG with CHARGE-AF Risk Score, there was no significant difference between the cumulative incidence of AF in the top quartile of the two methods^34^ whereas in our study the top percentiles of AI-estimated LA volume detected a significantly higher percentage of AF versus CHARGE-AF. Perhaps by directly identifying individuals with a very large LA volume, our approach is inherently more capable of detecting high-risk cases for future AF than other methods including ECG-based predictive AI models^36^.

### CAC Scans Can Provide More than CAC Scores

Our study corroborates findings from the Heinz Nixdorf Recall Study and others, and further brings to light the value of non-coronary findings in coronary calcium scans for a comprehensive CVD risk assessment beyond coronary heart disease^12,13,14,15^. Although manual and automated LA volumetry in chest CT scans are relatively novel^37,38^, the pathophysiology of enlarged LA and its relationship with AF is well understood^39,40^.

Several echocardiographic studies have shown that increased LA strain is associated with atrial arrhythmia^41,42,43,44^. Tsang et in 2001 reported that larger LA volume in echocardiographic studies was associated with a higher risk of AF in older patients. The predictive value of LA volume was incremental to that of clinical risk profile and conventional M-mode LA dimension^45,46^. Kizer et showed that LA size was an independent predictor of CVD events^26^.

Mahabadi et al^13^ showed in the longitudinal Heinz Nixdorf Recall Study that two-dimensional LA size and epicardial adipose tissue from non-contrast CT were strongly associated with prevalent and incident AF and that LA size diminished the link of epicardial adipose tissue with AF, and was also associated with incident major CV events independent of risk factors and CAC-score^14^.

In a study of 131 cases AutoChamber measurements in non-contrast cardiac CT scans were well correlated with automated cardiac chambers volumetry in contrast-enhanced cardiac CT scans using Philips Brilliance Workspace^47^. Similarly, AutoChamber measurements in 169 ECG-gated cardiac versus non-gated chest CT scans in the same patients (paired scans done same day) showed strong correlations (R^2^= 0.85-0.95 for different chambers)^48^.

### Limitations

Our study has some limitations. The MESA Exam 1 baseline CT scans, performed between 2000 and 2002, were predominantly conducted using electron-beam computed tomography (EBCT) scanners. This technology is no longer the commonly used method of CAC scanning. Since our AI training was done completely outside of MESA and used a modern multi-detector (256 slice) scanner, we do not anticipate this to affect the generalizability of our findings. Because MESA used the ICD codes to identify a history of AF at baseline and newly diagnosed AF, and it is known that ICD based diagnosis can be inaccurate (PPV 70–96%, median sensitivity 79%)^49^ it is likely that MESA missed some cases of AF.

## Conclusion

In this study, we presented AI-CAC data obtained from existing CAC scans in a large multi-ethnic prospective study and compared the predictive value of AI-CAC estimated LA volume versus the CHARGE-AF Score and BNP for predicting AF. AI-CAC LA volumetry enabled prediction of AF and improved on the predictive value of CHARGE-AF Risk Score and BNP.

## Clinical Perspectives

The potential value of non-coronary findings in coronary calcium scans is significant. The clinical utility of this opportunistic add-on to CAC scans warrants further validation in other longitudinal cohorts. Additionally, the high rate of AF in the 99^th^ percentile of AI-CAC LA volume makes it attractive for selection of participants into AF prevention clinical trials.

## Data Availability

All data produced in the present study are available upon reasonable request to the authors.

## Acknowledgements

Special thank you to Philip Greenland and Susan Heckbert for reviewing early versions of the manuscript.

This research was supported by 2R42AR070713 and R01HL146666 and MESA was supported by contracts 75N92020D00001, HHSN268201500003I, N01-HC-95159, 75N92020D00005, N01-HC-95160, 75N92020D00002, N01-HC-95161, 75N92020D00003, N01-HC-95162, 75N92020D00006, N01-HC-95163, 75N92020D00004, N01-HC-95164, 75N92020D00007, N01-HC-95165, N01-HC-95166, N01-HC-95167, N01-HC-95168 and N01-HC-95169 from the National Heart, Lung, and Blood Institute, and by grants UL1-TR-000040, UL1-TR-001079, and UL1-TR-001420 from the National Center for Advancing Translational Sciences (NCATS). The authors thank the other investigators, the staff, and the participants of the MESA study for their valuable contributions. A full list of participating MESA investigators and institutions can be found at http://www.mesa-nhlbi.org

## Funding and Acknowledgement

This research was supported by 2R42AR070713 and R01HL146666 and MESA was supported by contracts 75N92020D00001, HHSN268201500003I, N01-HC-95159, 75N92020D00005, N01-HC-95160, 75N92020D00002, N01-HC-95161, 75N92020D00003, N01-HC-95162, 75N92020D00006, N01-HC-95163, 75N92020D00004, N01-HC-95164, 75N92020D00007, N01-HC-95165, N01-HC-95166, N01-HC-95167, N01-HC-95168 and N01-HC-95169 from the National Heart, Lung, and Blood Institute, and by grants UL1-TR-000040, UL1-TR-001079, and UL1-TR-001420 from the National Center for Advancing Translational Sciences (NCATS).

